# Harmonization of SARS CoV-2 antibodies determination. Is it really possible?

**DOI:** 10.1101/2021.12.13.21267669

**Authors:** Ruggero Dittadi

## Abstract

The WHO standard was prepared with the aim of harmonizing assays detecting antibodies against SARS CoV-2. The aspect of the harmonization of the assays is to date under debate. We re-evaluated a previously studied set of cases (108 specimens of 48 patients and 60 specimens of 20 vaccinated subjects, collected after 14 days from the first dose, 14 days and 3 months after a second dose of the Comirnaty BNT162b2 vaccine), calculating the ratios between the results of two methods (SARS-CoV-2 IgG anti-RBD, SNIBE and anti-SARS-CoV-2 QuantiVac ELISA IgG, Euroimmun).

In the vaccinated subjects the ratios of the results between methods according to the WHO standard were relatively dispersed, but the harmonization results good. On the other hand, in patient samples the variability between tests was very high and the harmonization was unsatisfactory (median ratios between methods 2.23, 10th-90th percentile: 1.1-5.6).

Interestingly, in patient samples the harmonization depends on the time from the onset of symptoms, and greatly improves after 6 months from the diagnosis. 40 patient specimens and 31 of vaccinated subjects after the second dose were evaluated also with a third method (Access SARS-CoV-2 IgG (1st IS), Beckman Coulter), obtaining a similar trend.

We can conclude that the actual effectiveness of harmonization between methods may vary depending on the scenario in which they will be used.

---

At the end of 2020 the National Institute for Biological Standards and Control (NIBSC) established, using a pooled plasma obtained from individuals who recovered from COVID-19, the first WHO international standard for the immunoglobulins anti-SARS-CoV-2 (NIBSC 20/136). This standard could be used to calibrate in Binding Antibodies Units (BAU) the systems detecting antibodies against SARS CoV-2, in order to harmonize the different methods [1].

Few studies have so far directly evaluated the real effectiveness of this standard.

Perkmann et al [2] did not find significant harmonization in samples of vaccinated subjects after the first dose of BNT162b2, measured by five different methods. Lukaszuk et al [3] only marginally improved the comparison between two methods in vaccinated patients.

Infantino et al [4] evaluated different methods in a mix of patients and vaccinated subjects. They found on average a better comparability between the results but stated that the assays remain not interchangeable.

On the basis of the WHO standard, Ferrari et al [5] prepared another standard using only vaccinated subjects, with the aim of improving harmonization in this group of people. The results were encouraging but, although the correlations between methods were statistically significant, the distribution of cases remains rather dispersed.

A reason for the difficulty in harmonizing the methods certainly lies in the fact that the different methods are often built to detect antibodies directed towards different epitopes. However, the heterogeneity of antibodies produced at the onset of the disease may be a further cause of misalignment between different methods.

In a recently published study [6] we evaluated 108 specimens of 48 patients and 60 specimens of 20 vaccinated subjects, collected after 14 days from the first dose, 14 days and 3 months after a second dose of the Comirnaty BNT162b2 vaccine. We used a methods based on the determination of antibodies against receptor binding domain (SARS-CoV-2 IgG anti-RBD, SNIBE, Shenzen, China) performed on Maglumi platform and an ELISA method detecting antibodies against protein S1 (anti-SARS-CoV-2 QuantiVac ELISA IgG, Euroimmun, Lubeck, Germany).

We found that the transformation into Binding Antibodies Units allowed to harmonize the methods only in vaccinated subjects but not in the specimens of patients.

Our original data were re-evaluated calculating the ratios between the results of the two methods. Using the results expressed in the internal units of each method, the medians of the ratios between ELISA and Maglumi methods were 3.01 in patients, 1.89, 1.3 and 1.47 respectively in the vaccinated subjects after the first dose and 14 and 90 days after the second dose.

After the transformation into BAU/mL, the medians of the ratios between methods were 2.23 (10th-90th percentile: 1.1-5.6) in patients. In the vaccinated subjects, the ratios after the first dose (median 1.4 and 10th-90th percentile 0,93-2.1) were significantly higher (Kruskall-Wallis test: p=0.0009) than in the specimens 14 and 90 days after the second dose (overall median 0,98 and 10th-90th percentile 0.78-1.39).

The comparability between the two methods in vaccinated subjects after the second dose is good and the scattering is reasonably reduced. Less satisfactory was the comparability after the first dose. In the patients the differences between methods remain elevated even after the transformation in BAU and the dispersion of the ratios between the two methods was very high especially in the first weeks after the onset of the disease (up to about 10-15 weeks), to be reduced over time (fig 1). Interestingly, the 13 patient specimens collected more than 200 days after symptoms show a median ratio (1.1) and a variability (10th-90th percentile: 0.86-1.57) comparable to those of vaccinated subjects after the second dose. Probably the reduction of antibody heterogeneity over time affects the heterogeneity of antibody measurement by different methods. It is worth noting that in the 12 patients with more than one sample and at least one sample over 6 months after the symptoms’ onset, the decrease over time of the ratios between the two methods within each patient was evident, with a ratio between methods that tends progressively to 1 (fig.2).

**Figure 1.**
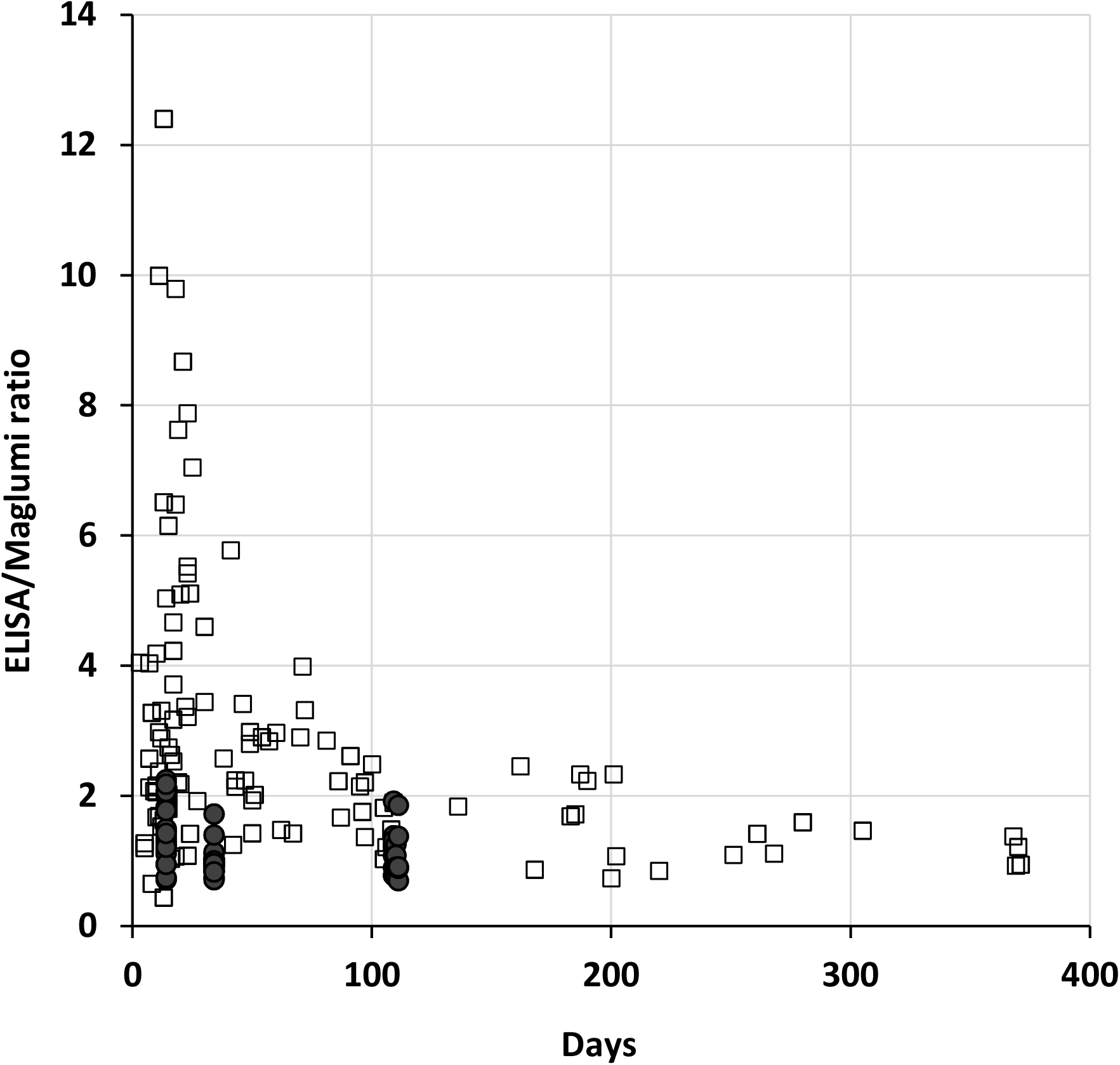
Plot of the ratios of each specimen between ELISA and Maglumi vs the days from the onset of symptoms, or from the first inoculum of vaccine. The open squares represent the patients specimens and the filled circles the vaccinated subjects.

**Figure 2.**
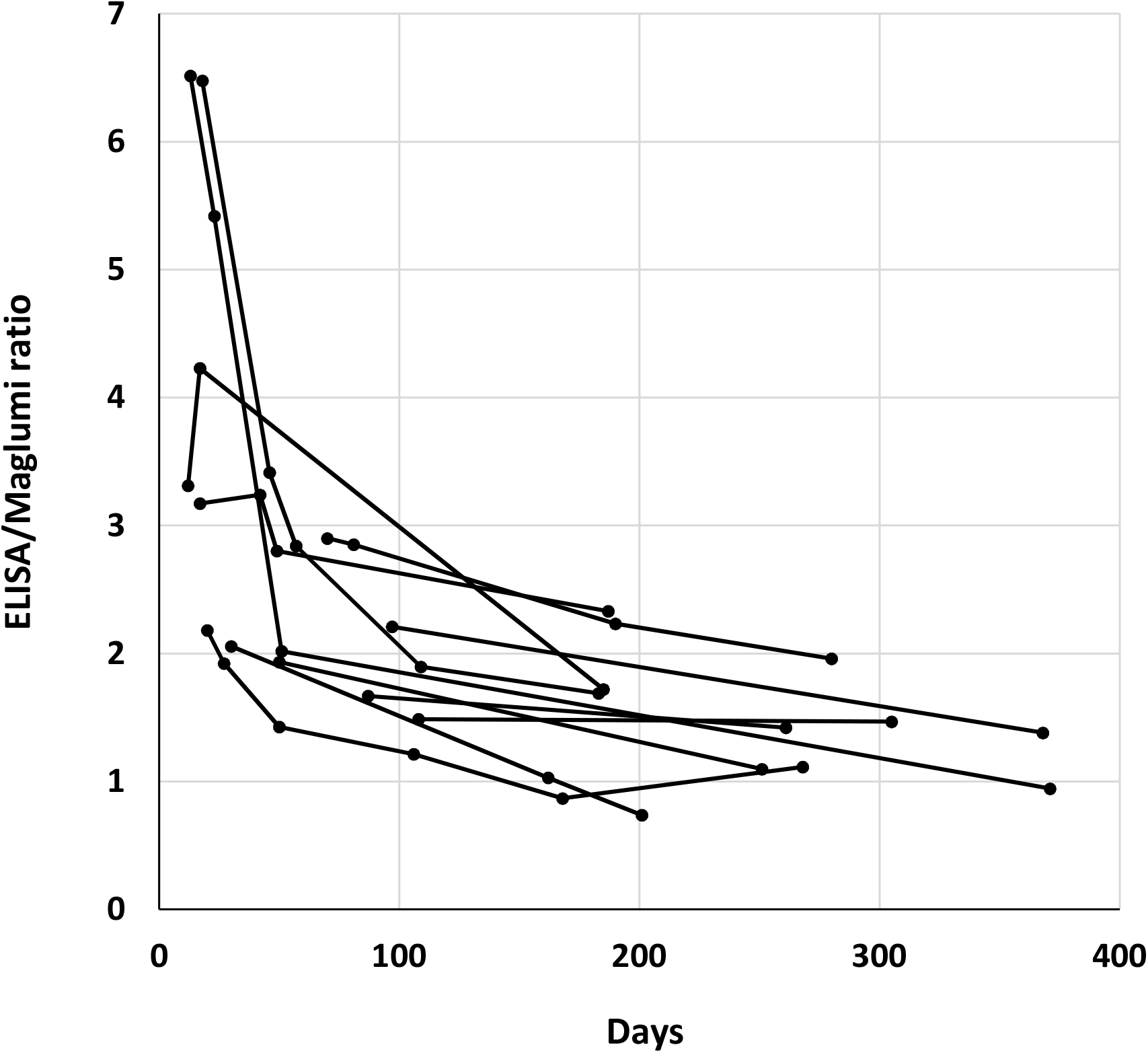
Plot of the ratios between ELISA and Maglumi vs the days from the onset of symptoms in the 12 patients with more than one sample and at least one sample over 6 months after the symptoms’ onset. Each patient was represented by a broken line.

A recently released Access SARS-CoV-2 IgG (1st IS) method, calibrated against WHO standard and carried out on the DxI 800 analyzer (Beckman Coulter, Inc. Brea, CA 92821 USA) was used to evaluate some of the cases previously studied (40 patient specimens and 31 of vaccinated subjects after the second dose), obtaining a similar behavior.

Median ratios between DxI and Maglumi were 1.96 (10th-90th percentile: 0.5-9.1) in patients and 0.62 (10th-90th percentile: 0.46-0.87) in vaccinated subjects. Median ratios between DxI and ELISA were 0.77 (10th-90th percentile: 0.35-2.18) in patients and 0.61 (10th-90th percentile: 0.35-0.88) in vaccinated subjects.

Despite the limited number of cases, it can be noted that the dispersion of the ratios between methods remains high in patients, and narrower in vaccinated subjects. However, harmonization is not very satisfactory. Even in vaccinated subjects, on average remains a difference of about 40% between DxI and the other two methods. Anyway, the decrease in the dispersion of the ratios between methods is confirmed when patients are examined after several months from the onset of the disease (Fig.3 and 4).

**Figure 3.**
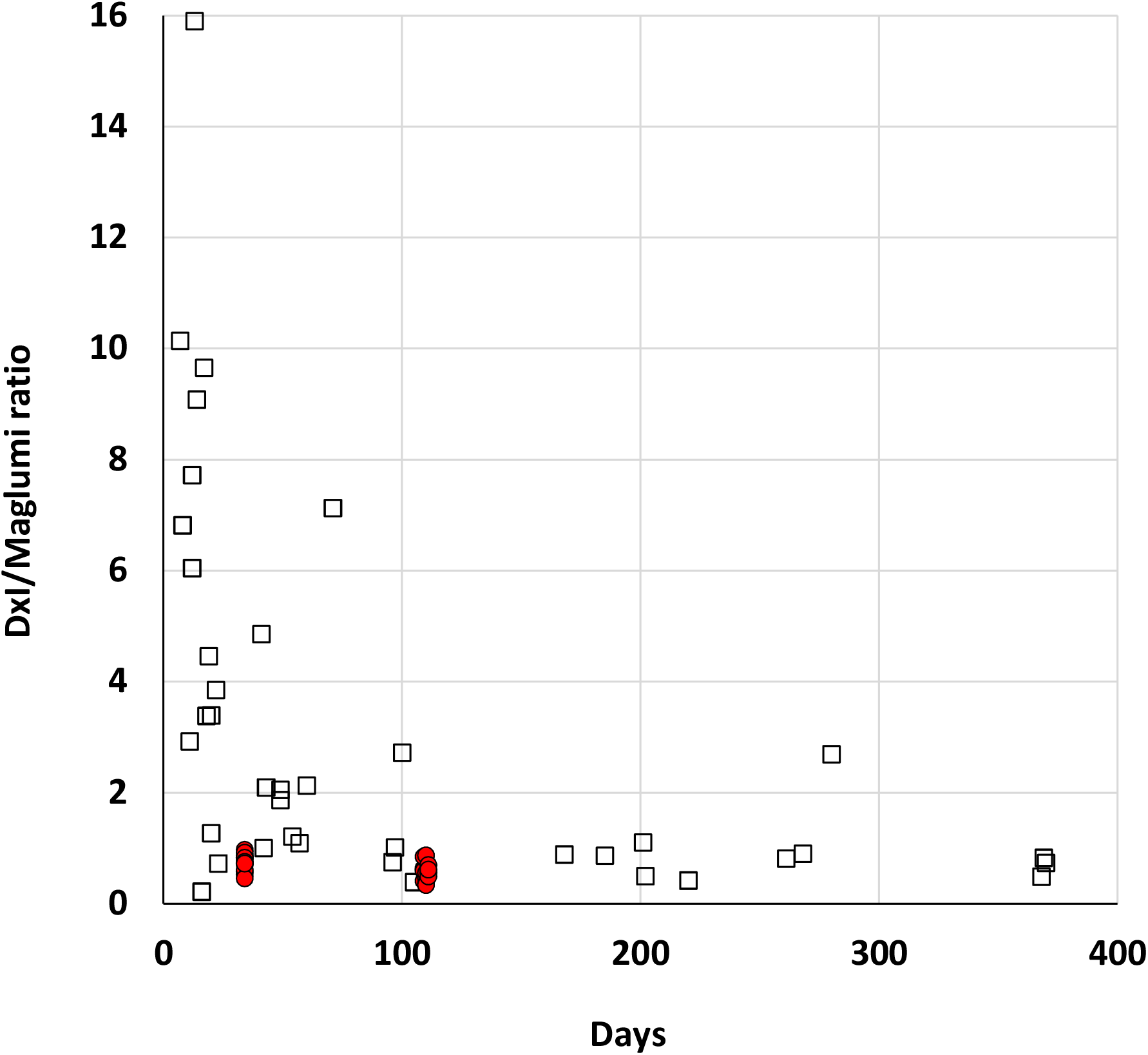
Plot of the ratios of each specimen between DxI and Maglumi vs the days from the onset of symptoms, or from the first inoculum of vaccine. The open squares represent the patients specimens and the red circles the vaccinated subjects.

**Figure 4.**
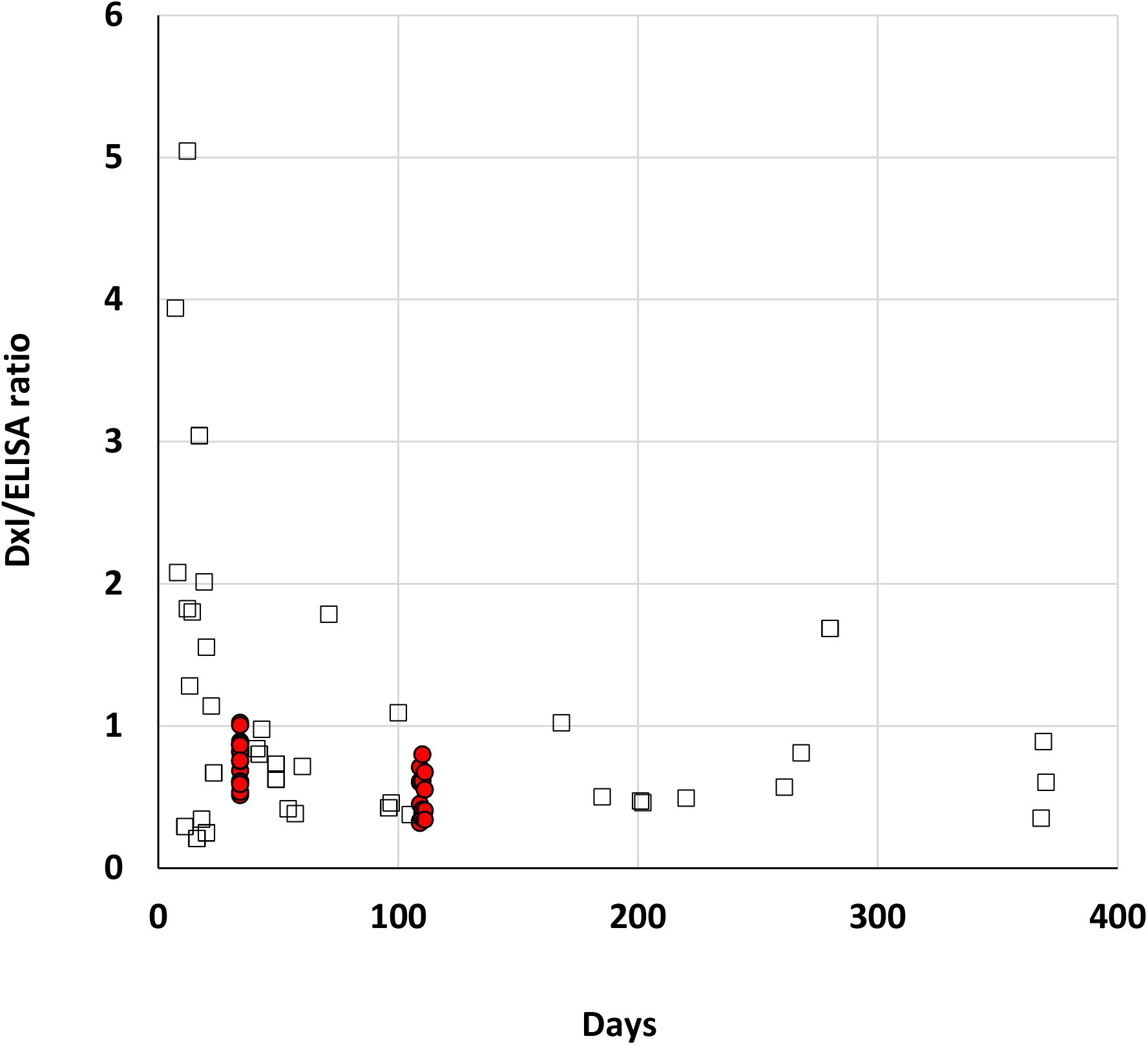
Plot of the ratios of each specimen between DxI and ELISA vs the days from the onset of symptoms, or from the first inoculum of vaccine. The open squares represent the patients specimens and the red circles the vaccinated subjects.

The evolution of antibodies favors the idiotypes against Receptor Binding Domain, probably making the antibody population more homogeneous [7] and allowing a better harmonization between methods by using the WHO standard.

The full realization of the harmonization seems rather complex and in certain scenarios may not be feasible.

For example, in the context of the monoclonal antibodies therapies, which can be administered to patients at the onset of the disease, the possibility of dose-dependent efficacy on the basis of the levels of antibodies has been proposed [8]. However, at least according to the results described in this study, in this case the attempt at harmonization between methods could be unfeasible, in part due to the large variability between methods in the first months of the disease.

On the other hand, the harmonization between methods could perhaps show some efficacy in scenarios where the antibody population is more homogeneous, e.g., for monitoring the vaccinated subjects and for establishing a correlate of protection based on antibodies levels [9,10].

## Data Availability

All data produced in the present study are available upon reasonable request to the authors

## Acknowledgments

The author would like to thank Euroimmun, Medical Systems S.p.A and Beckman Coulter for kindly providing the necessary reagents without any influence in study design and data analysis.

## Institutional Review Board Statement

The study was conducted according to the guidelines of the Declaration of Helsinki. The Ethical committee for clinical trials, ULSS3 Serenissima, Venice, approved the study (Approval n.149/A CESC).

## REFERENCES

1. Mattiuzzo P, Bentley EM, Hassal M, Routley S, Richardson S, Bernasconi V, Kristiansen P, Harvala H, Roberts D, Semple MG, Turtle LCW, Openshaw PJM, Baillie K. Establishment of the WHO International Standard and Reference Panel for Anti-SARS-CoV-2 Antibody; WHO Expert Committee on Biological Standardization: Geneva, Switzerland (2020) WHO/BS/2020.2403.

2. Perkmann T, Perkmann-Nagele N, Koller T, Mucher P, Marculescu R, Wolzt M, Wagner OF, Binder CJ, Haslacher H. Anti-Spike Protein Assays to Determine Post-Vaccination Antibody Levels: A Head-to-Head Comparison of Five Quantitative Assays. Microbiol Spectr 9 (2021) e0024721. doi: 10.1128/Spectrum.00247-21

3. Lukaszuk K, Kiewisz J, Rozanska K, Dabrowska M, Podolak A, Jakiel G, Woclawek-Potocka I, Lukaszuk A, Rabalski L. Usefulness of IVD Kits for the Assessment of SARS-CoV-2 Antibodies to Evaluate the Humoral Response to Vaccination. Vaccines 9 (2021) 840. doi: 10.3390/vaccines9080840

4. Infantino M, Pieri M, Nuccetelli M, Grossi V, Lari B, Tomassetti F, Calugi G, Pancani S, Benucci M, Casprini P, Manfredi M, Bernardini S. The WHO International Standard for COVID-19 serological tests: towards harmonization of anti-spike assays. Int Immunopharmacol 100 (2021) 108095. doi: 10.1016/j.intimp.2021.108095

5. Ferrari D, Mangia A, Spanò MS, Zaffarano L, Viganò M, Di Resta C, Locatelli M, Ciceri F, De Vecchi E. Quantitative serological evaluation as a valuable tool in the COVID-19 vaccination campaign. Clin Chem Lab Med. 59 (2021) 2019–2026. doi: 10.1515/cclm-2021-0364

6. Dittadi R, Seguso M, Bertoli I, Afshar H, Carraro P. Antibodies against SARS-CoV-2 Time Course in Patients and Vaccinated Subjects: An Evaluation of the Harmonization of Two Different Methods. Diagnostics (Basel) 11 (2021) 1709. doi: 10.3390/diagnostics11091709.

7. Wang Z, Muecksch F, Schaefer-Babajew D, Finkin S, Viant C, Gaebler C, Hoffmann HH, Barnes CO, Cipolla M, Ramos V, Oliveira TY, Cho A, Schmidt F, Da Silva J, Bednarski E, Aguado L, Yee J, Daga M, Turroja M, Millard KG, Jankovic M, Gazumyan A, Zhao Z, Rice CM, Bieniasz PD, Caskey M, Hatziioannou T, Nussenzweig MC. Naturally enhanced neutralizing breadth against SARS-CoV-2 one year after infection. Nature 595 (2021) 426–431. doi: 10.1038/s41586-021-03696-9.

8. Libster R., Perez Marc G., Wappner D, Coviello S, Bianchi A, Braem V, Esteban I, Caballero MT, Wood C, Berrueta M, Rondan A, Lescano G, Cruz P, Ritou Y, Fernández Viña V, Álvarez Paggi D, Esperante S, Ferreti A, Ofman G, Ciganda Á, Rodriguez R, Lantos J, Valentini R, Itcovici N, Hintze A, Oyarvide ML, Etchegaray C, Neira A, Name I, Alfonso J, López Castelo R, Caruso G, Rapelius S, Alvez F, Etchenique F, Dimase F, Alvarez D, Aranda SS, Sánchez Yanotti C, De Luca J, Jares Baglivo S, Laudanno S, Nowogrodzki F, Larrea R, Silveyra M, Leberzstein G, Debonis A, Molinos J, González M, Perez E, Kreplak N, Pastor Argüello S, Gibbons L, Althabe F, Bergel E, Polack FP. Early High-Titer Plasma Therapy to Prevent Severe Covid-19 in Older Adults. N Engl J Med 384 (2021) 610–618.

9. Earle KA, Ambrosino DM, Fiore-Gartland A, Goldblatt D, Gilbert PB, Siber GR, Dull P, Plotkin SA. Evidence for antibody as a protective correlate for COVID-19 vaccines. Vaccine 39 (2021) 4423–4428. doi: 10.1016/j.vaccine.2021.05.063.

10. Gilbert PB, Montefiori DC, McDermott AB, Fong Y, Benkeser D, Deng W, Zhou H, Houchens CR, Martins K, Jayashankar L, Castellino F, Flach B, Lin BC, O’Connell S, McDanal C, Eaton A, Sarzotti-Kelsoe M, Lu Y, Yu C, Borate B, van der Laan LWP, Hejazi NS, Huynh C, Miller J, El Sahly HM, Baden LR, Baron M, De La Cruz L, Gay C, Kalams S, Kelley CF, Andrasik MP, Kublin JG, Corey L, Neuzil KM, Carpp LN, Pajon R, Follmann D, Donis RO, Koup RA. Immune correlates analysis of the mRNA-1273 COVID-19 vaccine efficacy clinical trial Science (2021) eab3435. Online ahead of print.

